# Using machine learning and clinical registry data to uncover variation in clinical decision making

**DOI:** 10.1101/2022.10.06.22280684

**Authors:** Charlotte James, Michael Allen, Martin James, Richard Everson

## Abstract

Clinical registry data contains a wealth of information on patients, clinical practice, outcomes and interventions. Machine learning algorithms are able to learn complex patterns from data. We present methods for using machine learning with clinical registry data to carry out retrospective audit of clinical practice. Using a registry of stroke patients, we demonstrate how machine learning can be used to: investigate whether patients would have been treated differently had they attended a different hospital; group hospitals according to clinical decision making practice; identify where there is variation in decision making between hospitals; characterise patients that hospitals find it hard to agree on how to treat. Our methods should be applicable to any clinical registry and any machine learning algorithm to investigate the extent to which clinical practice is standardized and identify areas for improvement at a hospital level.

## 1. Introduction

Clinical registries are collections of information on people that have a specific disease, have experienced, or are at-risk of, an adverse health-related event or have been exposed to something that might have an adverse effect. They provide information on clinical practice, patient outcomes and the effect of interventions and are therefore useful for monitoring the course of disease, examining the effect of a treatment on patient outcome and measuring quality of care [1].

In England the Healthcare Quality Improvement Partnership (HQIP), on behalf of the National Health Service (NHS), is responsible for over-seeing and commissioning more than 30 clinical audits (registries), which form the National Clinical Audit Programme. These collect and analyse registry data supplied by clinicians and are intended to be used for quality improvement by allowing the quality of care and services to be measured against agreed standards and for improvements to be made where necessary [2]. The quantity and quality of data contained in these registries has the potential to provide insight into additional processes that can have an impact on patient outcome, such as clinical decision making. As decision making processes are complex and involve many interacting factors, standard statistical analyses are unlikely to capture this insight.

Machine learning algorithms are able to learn complex patterns from data. They have shown promise in many areas, including drug discovery [3], precision medicine [4], medical diagnosis [5, 6], and clinical decision support [7, 8] suggesting that, given the right data, ML algorithms can learn clinical decision making processes. As the data contained in clinical registries includes information on patients, clinical practice, treatment and outcomes, ML presents an opportunity to understand the decision making processes of clinicians, assess the extent to which these processes are standardized and, if not, uncover where decision making varies.

The national audit covering stroke is the Sentinel Stroke National Audit Programme (SSNAP) [9]. SSNAP collects longitudinal data on the processes and outcomes of stroke care up to 6 months post-stroke for more than 95% of emergency stroke admissions to acute hospitals in England, Wales and Northern Ireland. Every year data from approximately 85,000 patients are collected. SSNAP publishes quarterly and yearly analysis of results on its website. Data from SSNAP has been previously used to investigate clinical decision making [10], socioeconomic risk factors for stroke and their influence on care received and patient outcome [11], and temporal variation in stroke care [12].

Stroke is a leading cause of death and disability worldwide which occurs when there is a blood clot or bleed in the brain preventing blood from reaching parts of the brain. Around 80-85% of strokes are due to a blood clot (ischemic stroke). Thrombolysis is the only licensed drug treatment for acute ischemic stroke. It is critically time dependent: the longer the time window between stroke and administration, the less benefit to the patient. After 4.5 hours there is little or no benefit of thrombolysis [13]. In England, Wales and Northern Ireland, 11.1% of patients with confirmed acute stroke receive thrombolysis, however there is considerable variation between hospitals, from 0% to 24.5%. Reasons for this variation are known to include slow uptake of the treatment and in-hospital delays to administration [14]; it is also possible that differences in clinical decision making play a part [15].

To investigate whether variation in thrombolysis rates between hospitals is due, at least in part, to differences in clinical decision making we used a ML approach. Specifically, we trained ML algorithms to learn the decision making outcome in each hospital. The algorithms are trained using data that is available to the clinician and that the clinician would use to decide whether or not to administer thrombolysis to a patient. The algorithms predict the binary outcome, to thrombolyse (1) or not (0). By using ML to learn the decision making processes in a hospital, we are able to answer counterfactual, or ‘what if?’, questions, such as ‘what if a patient had been admitted to a different hospital?’. This allows us to determine whether differences in clinical decision making contribute to the variation in thrombolysis rates observed between hospitals, and characterize patients that hospitals find it harder to agree how to treat.

While we focus on clinical decision making surrounding the use of thrombolysis for acute stroke, the methods we present should be applicable to any clinical registry data. These methods could be used with clinical registries to retrospectively assess variation in decision making between hospitals, group hospitals according to their decision making and characterise patients that clinicians find it difficult to agree how to treat. Uncovering differences in clinical practice using these methods may provide an opportunity to improve patient care by standardizing treatment across the board.

## 2. Materials & Methods

### 2.1. Data

We used data from SSNAP collected over 3 years, from January 2016 to December 2018, corresponding to 246,676 emergency stroke admissions admitted to 180 acute care hospitals. We restricted our analysis to hospitals that admitted at least 100 patients during the 3 year time period and thrombolysed 10 or more. As thrombolysis is only considered to have a significant effect if given within 4.5 hours of stroke onset [13] and we are interested in emergency stroke admissions, only patients that had a stroke out of hospital and arrived in hospital within 4 hours of known stroke onset were included in the analysis. This resulted in 132 unique hospitals and 88,928 stroke patients, of which 26,257 (29.5%) received thrombolysis.

The data contained 47 features: admitting hospital (N=1), patient characteristics (N=5), pathway information (N=10), comorbidities (N=10), National Institutes of Health Stroke Scale (NIHSS) (N=16), other clinical information (N=4) and whether thrombolysis was given (N=1) (Table S1). The NIHSS is a 15 item neurological scale that is used by clinicians as a measure of stroke severity. It ranges from 0 to 42 and may be used to classify a stroke as mild (NIHSS 1-4), moderate (NIHSS 5-14), severe (NIHSS 15-24) or very severe (NIHSS > 25) [16]. Also present in the data is a patient’s modified Rankin Scale (mRS) score before stroke, which is a measure of pre-stroke disability ranging from 0 to 5, with 0 corresponding to no prior disability and 5 meaning severely disabled.

Of the 47 features, 24 were missing for at least one patient (Table S2). If components of the NIHSS were missing they were replaced with 0 and if the time from arrival to brain scan (*Brain Imaging Time (minutes)*) was missing it was replaced with 9999, both of which force the probability of thrombolysis to be low. If a missing feature was binary it was coded as ‘missing’. All binary and categorical variables were one-hot-encoded, with ‘missing’ treated as a separate category.

### 2.2. Learning clinical decision making

We used a ML approach to learn clinical decision making outcomes from the data.

Specifically, we implemented a Random Forest (RF) which is an example of an ensemble learning algorithm composed of individual decision trees. We used RF as it is known to be robust and have a lower variance compared to a single decision tree. During training a RF will select a random sample of the training data with replacement and fit a decision tree to the sample. This process is repeated many times, the exact number being a parameter of the algorithm, to create an ensemble (or forest) of decision trees [17].

The RF is trained to perform a binary classification task: to decide whether a patient should be thrombolysed (class 1) or not (class 0). Each decision tree in the RF performs this classification task independently. A patient’s final class is determined by averaging over the outcomes of all decision trees: if the average is less than the decision threshold, the patient is assigned class 0 (not thrombolysed) else the patient is assigned class 1 (thrombolysed).

To implement the RF, we used Python’s sci-kit learn library [18] incorporating stratified 10-fold cross-validation: the data was split into a training set (90%) and validation set (10%) ten times, such that each patient is in the validation set exactly once. The RF was fit to the training set at each fold and the validation set was used to assess performance. Final performance measures were determined by averaging over those for each fold.

### 2.3. Hospital models

We divided our data into separate hospitals and trained an RF on patients from each hospital. Each of these RFs learn the decision making within a single hospital during training. We again used stratified 10-fold cross-validation and assessed how well each model was able to learn the decision making at each hospital. The decision threshold of each model was calibrated such that the predicted positive rate (i.e. fraction of patients predicted to be given thrombolysis) was equal to the true positive rate.

### 2.4. Assessing variation in decision making

To investigate how decision making varies between hospitals, we again trained RFs on patients from each hospital, using all patients from that hospital for training, as opposed to 10-fold cross validation. We subsequently took all other patients in our data set and applied each hospital’s RF to every patient: the classification given by each RF represents what would have happened if a patient had been treated at that hospital.

### 2.5. Grouping hospitals by decision making

To find hospitals with similar decision making processes, we set aside a representative cohort of 10,000 patients. We trained an RF on patients from each hospital selected from the remaining 78,928 patients. Each RF was applied to the cohort of patients to determine whether or not each patient would have been thromoblysed had they attended each hospital. We computed the pairwise Hamming distance, *D*_*H*_, between the predicted outcomes for every pair of hospitals. The Hamming distance is the proportion of patients on which the two hospital models disagree on the outcome: if *D*_*H*_ is close to 0 the hospitals would make similar decisions, whereas if it is close to 1 their modelled decision making processes are different.

## 3. Results

### 3.1. Population characteristics

Population characteristics are displayed in Table 1. Patients that were thrombolysed were, on average, younger (mean 73 years), arrived at the hospital in less time (mean 97 mins), had brain imaging scans faster (mean 23 mins), had more severe stroke (mean NIHSS 11.6) and lower levels of disability (mean rankin 0.7) compared to those that were not thrombolysed.

**Table 1:** Population characteristics stratified by outcome for patients in the treatment pathway. ^1^Values correspond to mean (std) unless otherwise stated. ^2^Standard deviation affected by imputation method

| Measure<sup>1</sup> | Thrombolysed N = 26257 | Not Thrombolysed N = 62671 |
| --- | --- | --- |
| Age (years) | 73 (14) | 76 (14) |
| Female (N, %) | 12084 (46) | 30796 (49) |
| White (N, %) | 22861 (87) | 55660 (89) |
| Onset to Arrival (minutes) | 97 (46) | 117 (54) |
| Arrival to Imaging (minutes) | 23 (20) | 157 (1095)<sup>2</sup> |
| NIHSS | 11.6 (7.0) | 8.4 (8.4) |
| Facial Palsy | 1.2 (0.9) | 0.8 (0.8) |
| Dysarthria | 1.0 (0.8) | 0.7 (0.8) |
| Best Language | 1.1 (1.1) | 0.7 (1.1) |
| Rankin before Stroke | 0.7 (1.2) | 1.2 (1.5) |

### 3.2. Learning clinical decision making

Model performance is described in Table 2: the RF trained on all patients performed well with a mean accuracy of 84.3% (range 83.6 - 84.7) and AUC of 0.91 (range 0.91 - 0.92). An accuracy of 85% means that 15% of patients were misclassified (Combined Model, Table 2). If clinical decision making was consistent both within and between hospitals we would expect misclassified patients to lie close to the decision boundary: these patients represent borderline decisions. To verify this, we looked at the probability distribution for misclassified patients, where the probability is the average outcome over all trees in the RF. From Figure 1 it is clear that while the distribution is approximately centred on the decision threshold there is a significant proportion of patients who were confidently misclassified: the probability of receiving thrombolysis is close to 0 or 1. This shows that not all misclassified patients represent borderline decisions and therefore supports our hypothesis that there is variation in clinical decision making: a confident misclassification means that there was a similar patient in the training data who was treated differently.

**Table 2:** Performance measures of Random Forest algorithm trained on data from all hospitals together with one-hot encoding of hospital ID (Combined Model) and each hospital separately (Hospital Models) ^1^For a decision threshold of 0.5. ^2^Values correspond to mean (range) of 10 folds ^3^Values correspond to mean (range) of all 132 hospital models. ^4^ AUC independent of decision threshold

| Performance Measure | Combined Model<sup>1,2</sup> | Hospital Models<sup>3</sup> |
| --- | --- | --- |
| Accuracy | 84.3 (83.6 - 84.7) | 84.5 (76.1 - 95.2) |
| Sensitivity | 0.69 (0.67 - 0.70) | 0.72 (0.43 - 0.86) |
| Specificity | 0.91 (0.90 - 0.91) | 0.86 (0.76 - 0.97) |
| False positive rate | 0.09 (0.09 - 0.10) | 0.11 (0.03 - 0.24) |
| False negative rate | 0.31 (0.30 - 0.33) | 0.28 (0.14 - 0.56) |
| Positive predictive value | 0.69 (0.67 - 0.70) | 0.72 (0.43 - 0.86) |
| Negative predictive value | 0.91 (0.90 - 0.91) | 0.89 (0.76 - 0.97) |
| Positive rate | 0.27 (0.26 - 0.27) | 0.29 (0.06 - 0.50) |
| AUC<sup>4</sup> | 0.91 (0.91 - 0.92) | 0.90 (0.80 - 0.97) |

**Figure 1:**
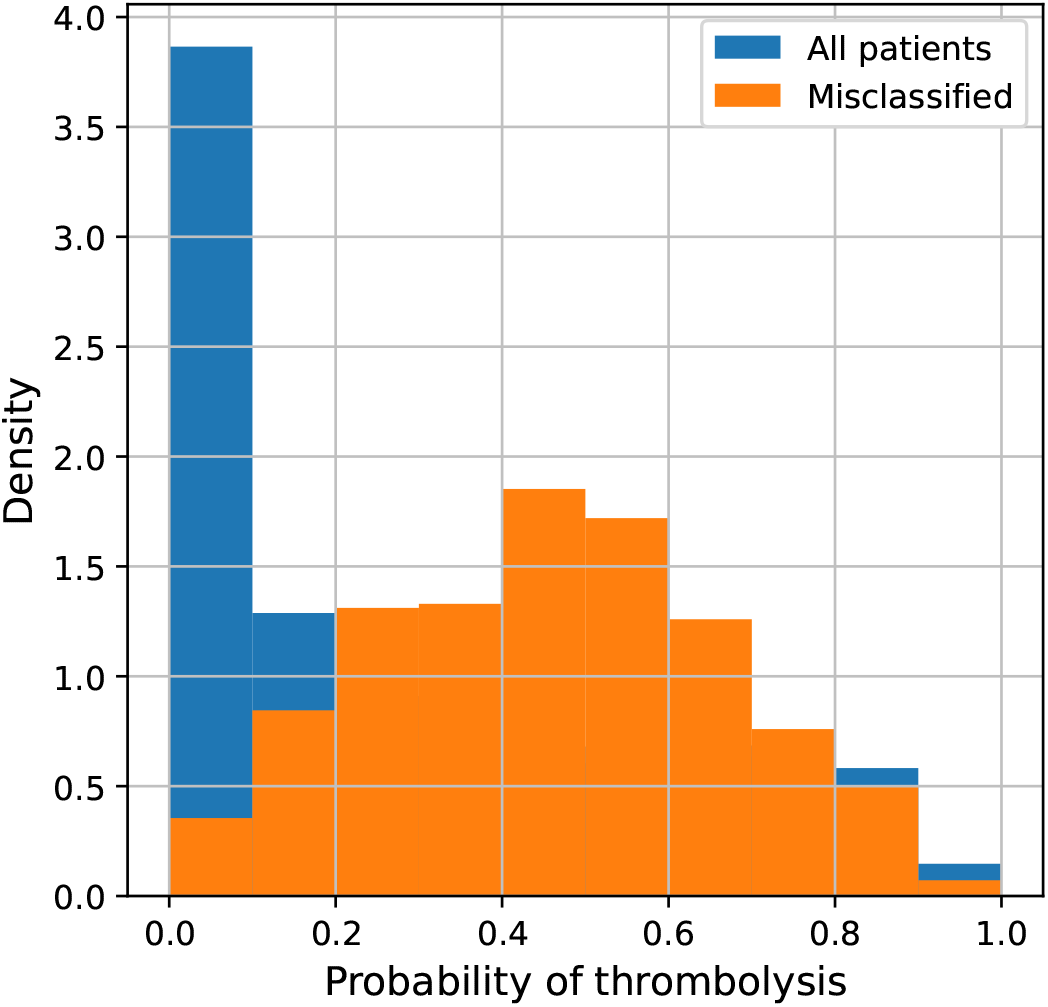
Probability distribution for patients misclassified by a Random Forest algorithm trained on data from all hospitals together.

### 3.3. Hospital models

In order to understand sources of variation in decision making between hospitals, we trained separate RFs on the patients from each of the 132 hospitals. On average, individual hospital models perform similarly to the combined model (Table 2). However, for each measure there is a range of values: the lowest performing hospital model has an accuracy of 76.1% and AUC of 0.80, which is significantly lower than the accuracy and AUC of the combined model (84.3% and 0.91). The range of values of the performance measures may be explained by variation in hospital size and thrombolysis rates. The performance of each hospital model will be affected by both the amount of data (hospital size) and class balance (thrombolysis rate): hospitals with a smaller number of patients and lower thrombolysis rate are less likely to have well calibrated models.

We assessed how well hospital models were calibrated by plotting a calibration curve for each model (Figure S1a). If a model is perfectly calibrated, the proportion of a set of patients that are assigned an equal probability of receiving thrombolysis by the model, that go on to receive thrombolysis, should be equal to the probability: i.e. out of all patients the model assigns a 60% chance of receiving thrombolysis to, 60% of the patients should receive it.

From Figure S1a, most hospital models are well calibrated, however some do not output probabilities over the whole range, or are not well calibrated at higher probabilities (Figure S1b). In order to improve model calibration we applied Platt Scaling [19], fitting a logistic regression to the probabilities output by each hospital model. After applying Platt Scaling, the model calibration curves are closer to that of the perfectly calibrated model (black dashed line, Figure S1c). Figure S2 shows how hospitals with models that were not well calibrated compare to other hospitals in terms of number of patients and thrombolysis rate. From this it is clear that hospitals that have both fewer patients and lower thrombolysis rates are less likely to be well calibrated. This may be attributed to the training data for these hospital models being both smaller in size and more imbalanced.

As with the combined model, hospital models also confidently misclassify patients (Figure S3). In this case, confident misclassifications represent cases were a patient was treated differently to what would be expected in that hospital: for a confident misclassification to occur, a similar patient that was treated differently must be present in the training data. However, the reason that these patients were treated differently to the norm for the hospital they attended is likely not captured in the data and will not be explored further.

### 3.4. Assessing variation in decision making

Using hospital models, each trained on all patients from a single hospital, we determined how each patient in the dataset would have been treated had they attended a different hospital. We found that it is much easier for hospitals to agree who not to thrombolyse than who to give thromobolysis to (Figure 2). Specifically 80% of hospitals agree not to thrombolyse 85% of patients that did not receive thrombolysis, where as only 60% of patients who did receive thrombolysis would have received it at 80% of hospitals. Only 3% of patients that were given thrombolysis would have received it at every hospital.

**Figure 2:**
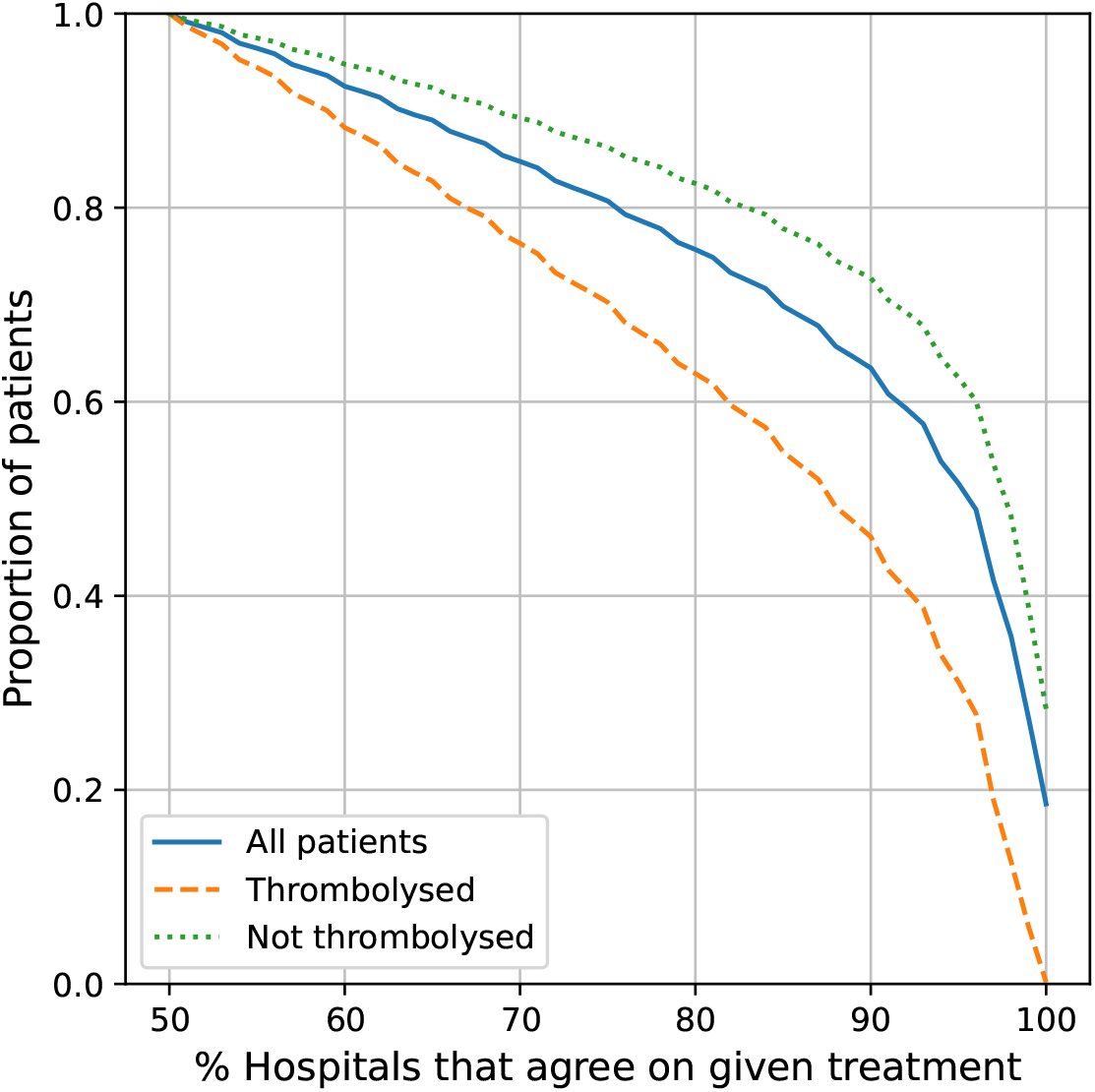
Proportion of patients that hospitals agree to treat, stratified by patient outcome.

### 3.5. Grouping hospitals by decision making

To understand where the variation in decision making occurs, and to group hospitals according to their decision making, we set aside a representative cohort of 10,000 patients and re-trained hospital models on the remaining 78,928 patients. Computing *D*_*H*_ between the predicted outcomes of each pair of hospitals we found no clear groups of hospitals (Figure S4), due to the fact that hospitals find it easy to agree on who not to thrombolyse.

Accounting for this, we re-calculated *D*_*H*_ for each pair of hospitals using only *contentious* patients: patients that would have been thrombolysed at 30-70% of hospitals, of which there were 1,364, reasoning that it is these borderline cases that hospitals with similar decision making processes would make similar decisions on. We found two distinct groups of hospitals (Figure 3). Looking at the thrombolysis rate in contentious patients for each of these groups, the largest group (location 108-118 in Figure 3) has a high thrombolysis rate in this group of patients, with a mean of 86%, suggesting that these hospitals are more likely to make the decision to thrombolyse when others may consider it risky. In contrast, the smaller group in the top left of the figure has a low thrombolysis rate, treating on average 16% of contentious patients: these are hospitals that are less likely to take risks when the decision to thrombolyse is borderline.

**Figure 3:**
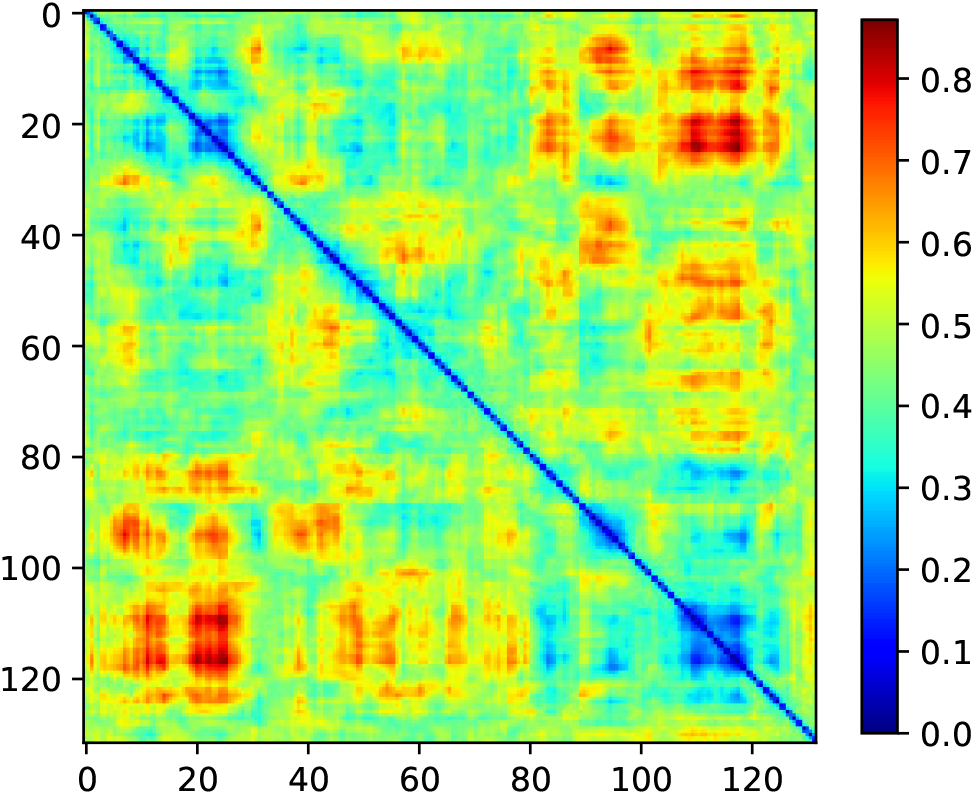
Pairwise Hamming distance, *D*_*H*_, between hospitals, derived from the decisions made at each hospital on how to treat a standardised cohort of 10,000 patients.

### 3.6. Characterising patients where decision making differs

We characterized the group of contentious patients by comparing them to patients that were not contentious: patients that over 70% of hospitals agreed on the treatment of. We found that contentious patients did not differ significantly in terms of age, arrival time, scan time or stroke severity (NIHSS) compared to patients that over 70% of hospitals would have thrombolysed (Table 3). However there was a difference in terms of disability: the modified rankin score of contentious patients was 1.1 *±* 1.4 where as for those patients that hospitals agreed to thrombolyse, this was 0.6 *±* 1.1, and for patients hospitals agreed not to thrombolyse it was 1.2 *±* 1.5. This suggests patients who have a prior disability represent borderline decisions in terms of whether or not thrombolysis should be given, and more severe disability reduces the chance of thrombolysis.

**Table 3:** Key measures for patients in a cohort of 10,000 that over 70% of hospitals would have thrombolysed (Thrombolysed), over 70% of hospitals wouldn’t have thrombolysed (Not Thrombolysed) and those that were contentious (30-70% of hospitals would have thrombolysed). ^1^Values correspond to mean (std).

| Measure<sup>1</sup> | Thrombolysed | Not Thrombolysed | Contentious |
| --- | --- | --- | --- |
| Age | 73 (14) | 76 (14) | 75 (14) |
| Onset to Arrival | 89 (41) | 123 (55) | 101 (47) |
| Arrival to Imaging | 19 (12) | 150 (890) | 26 (24) |
| NIHSS | 13 (7) | 6 (7) | 12 (8) |
| Rankin before Stroke | 0.6 (1.1) | 1.2 (1.5) | 1.1 (1.4) |

To further understand the difference between contentious patients and those that over 70% of hospitals would have thrombolysed, we trained an additional RF to classify patients into these groups. The RF again performs a binary classification task to determine whether a patient would have been thrombolysed (class 0) or was contentious (class 1). The performance of the RF is a measure of how easy it is to distinguish between these two groups of patients, and determining which features are important for the classification provides insight into how these groups of patients differ.

The RF was able to classify patients with an accuracy of 88% (decision threshold of 0.5) and an AUC of 0.96, suggesting that contentious patients are distinguishable from those that 70% of hospitals would have thrombolysed. To understand how these patients differ, we performed a principal component analysis by re-training the RF with an increasing number of features, starting with the most important. We found that the AUC of the classifier reaches 0.96 with less than 20 features (Figure 4). Of these features, the most important are NIHSS on arrival and whether the stroke onset time is known or an estimate. From Figure 5a it is clear that patients that are contentious are more likely to have an estimated onset time than those that hospitals agree to thrombolyse. Figure 5b shows, for each NIHSS score, the proportion of patients in each group. Here it is clear that hospitals agree not to thrombolyse patients with very mild stroke, where as contentious patients have moderate to severe stroke (NIHSS 5-30). This pattern is also clear from Figure 5c, where we compare the proportion of patients that hospitals agree how to treat (thrombolyse or not thrombolyse), to the contentious group for each NIHSS score. The number of patients with NIHSS *>* 30, the number of patients is very low (Figure 5d) therefore the distributions in Figures 5b and c at higher NIHSS values are less representative of the true distribution, as shown by the larger margin of uncertainty in Figure 5c.

**Figure 4:**
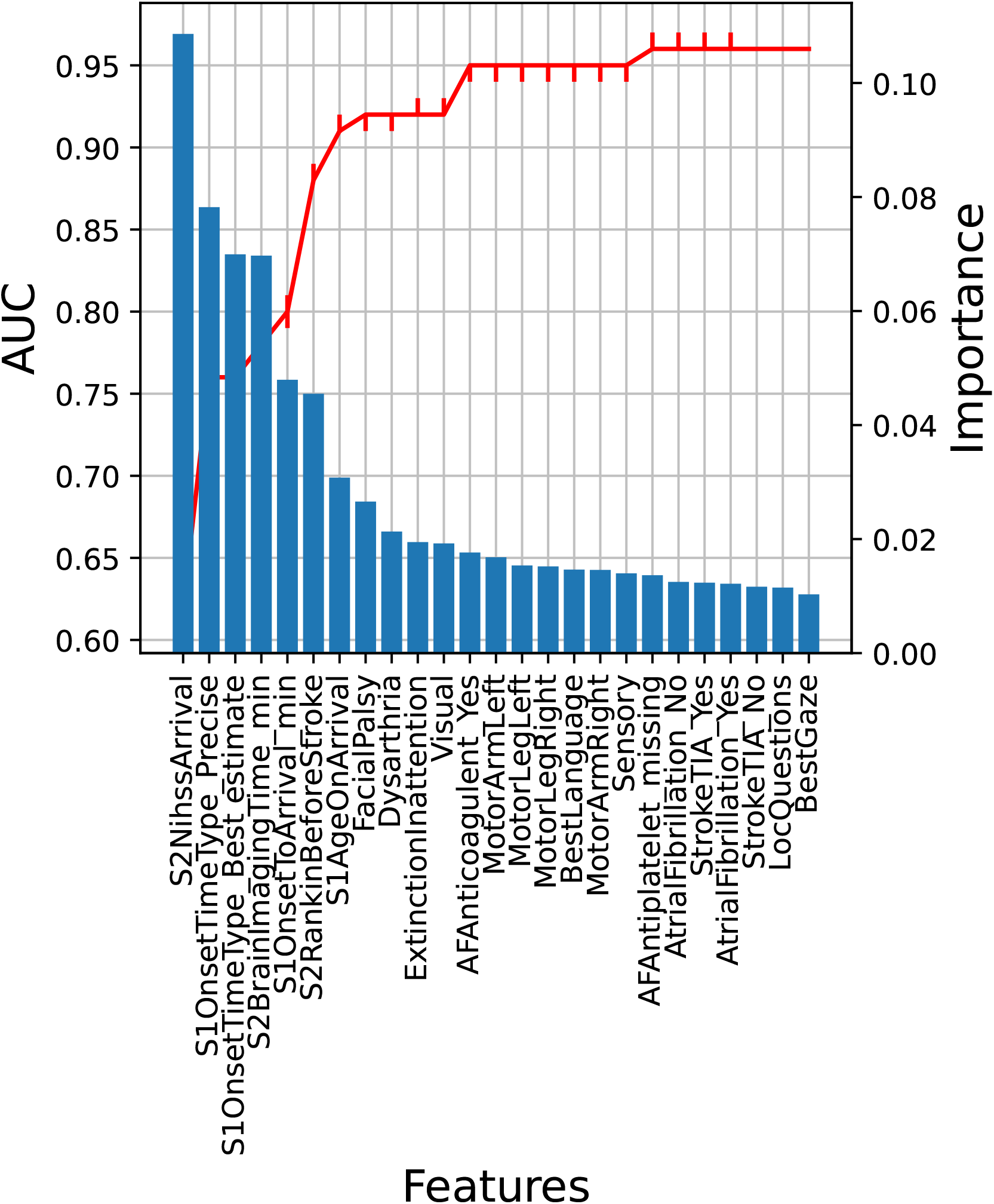
Features, ordered by importance (x-axis) vs importance (y-axis right) and cumulative AUC (y-axis left) for a random forest trained to classify whether hospitals would agree to thrombolyse a patient or whether they are contentious. Red line represents the median AUC and error bars correspond to 10th and 90th percentiles after bootstrapping the training data 100 times.

**Figure 5:**
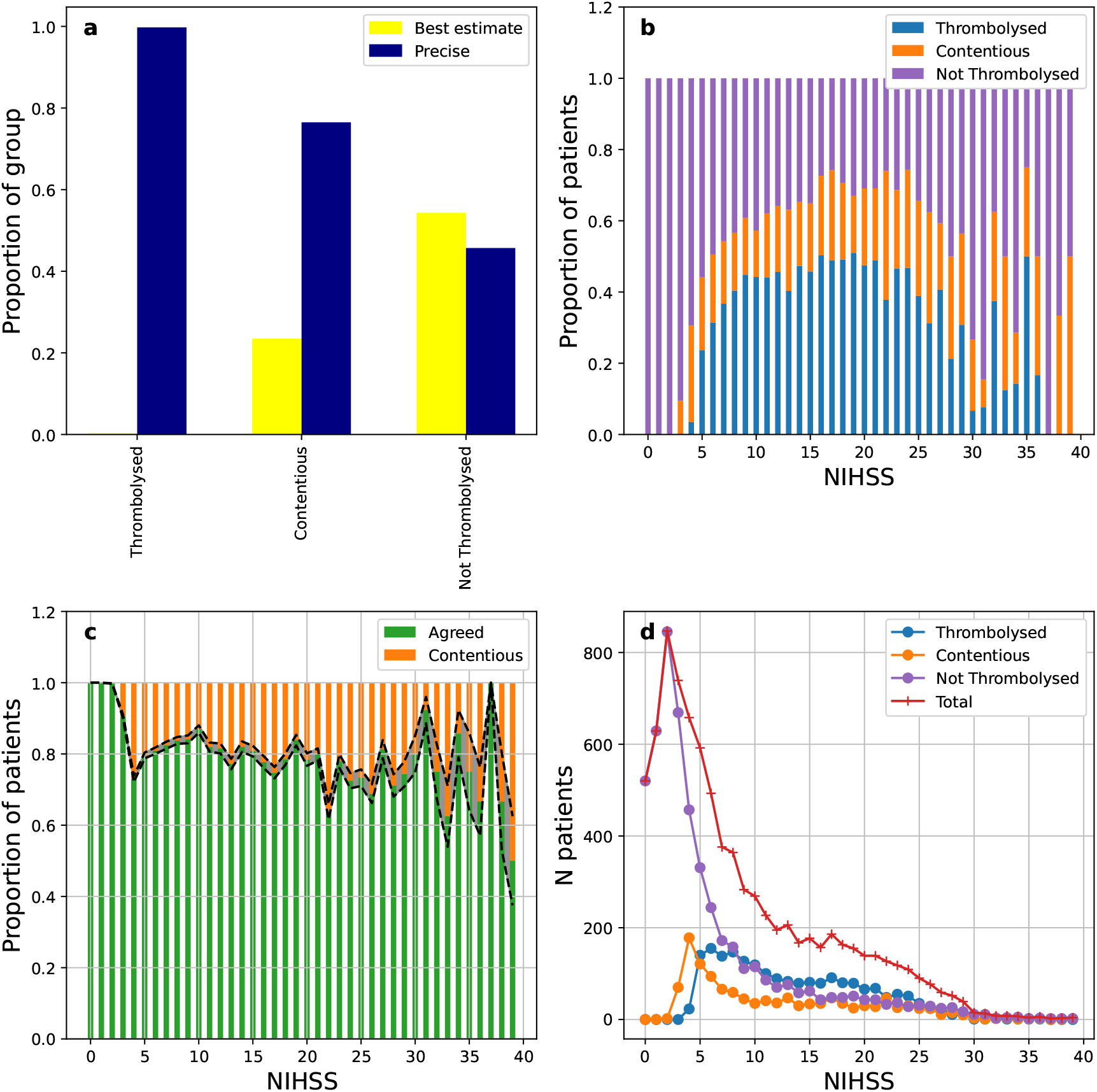
**a** Distribution of onset time types for patients in a cohort of 10,000 that over 70% of hospitals would have thrombolysed (Thrombolysed), patients that over 70% of hospitals wouldn’t have thrombolysed (Not Thrombolysed) and contentious patients. **b** Distribution of NIHSS values for patients in a cohort of 10,000 that over 70% of hospitals would have thrombolysed (Thrombolysed, blue), over 70% of hospitals wouldn’t have thrombolysed (Not Thrombolysed, purple) and contentious patients (orange). **c** Distribution of NIHSS values for patients that over 70% of hospitals agreed on how to treat (Agreed, green) and contentious patients (orange). **d** Distribution of NIHSS values for patients in a cohort of 10,000 (Total, red) and stratified by whether over 70% of hospitals would have thrombolysed (Thrombolysed, blue), over 70% of hospitals wouldn’t have thrombolysed (Not Thrombolysed, purple) and contentious patients (orange).

For the RF, feature importance is determined by the Gini importance: for each tree a feature’s importance is the total decrease in impurity that occurs when the feature is used to split a node, weighted by the number of samples the node splits. The Gini importance is calculated for each tree and then averaged to give a final feature importance. Shapley values are an alternative method for determining how each feature contributes to a classification. While Gini importance gives an overall measure of importance for each feature, Shapley values contain information on how the value of a feature contributes to the classifier’s final decision. Computing Shapley values themselves is computationally intense, however SHAP (SHapley Additive exPlanations) [20] values provide an estimation of Shapley values and can be used to explain how features contribute to model predictions.

Figure 6 displays the SHAP values for the 20 features that have the highest impact on the output of the RF trained to distinguish between patients that hospitals agree to thrombolyse and those that are contentious. For each feature on the y-axis, each point represents the SHAP value of that feature for a single patient. The SHAP value itself (x-axis) tells us whether the feature pushed the model towards classifying a patient as contentious (class 1, positive SHAP) or not (class 0, negative SHAP).

**Figure 6:**
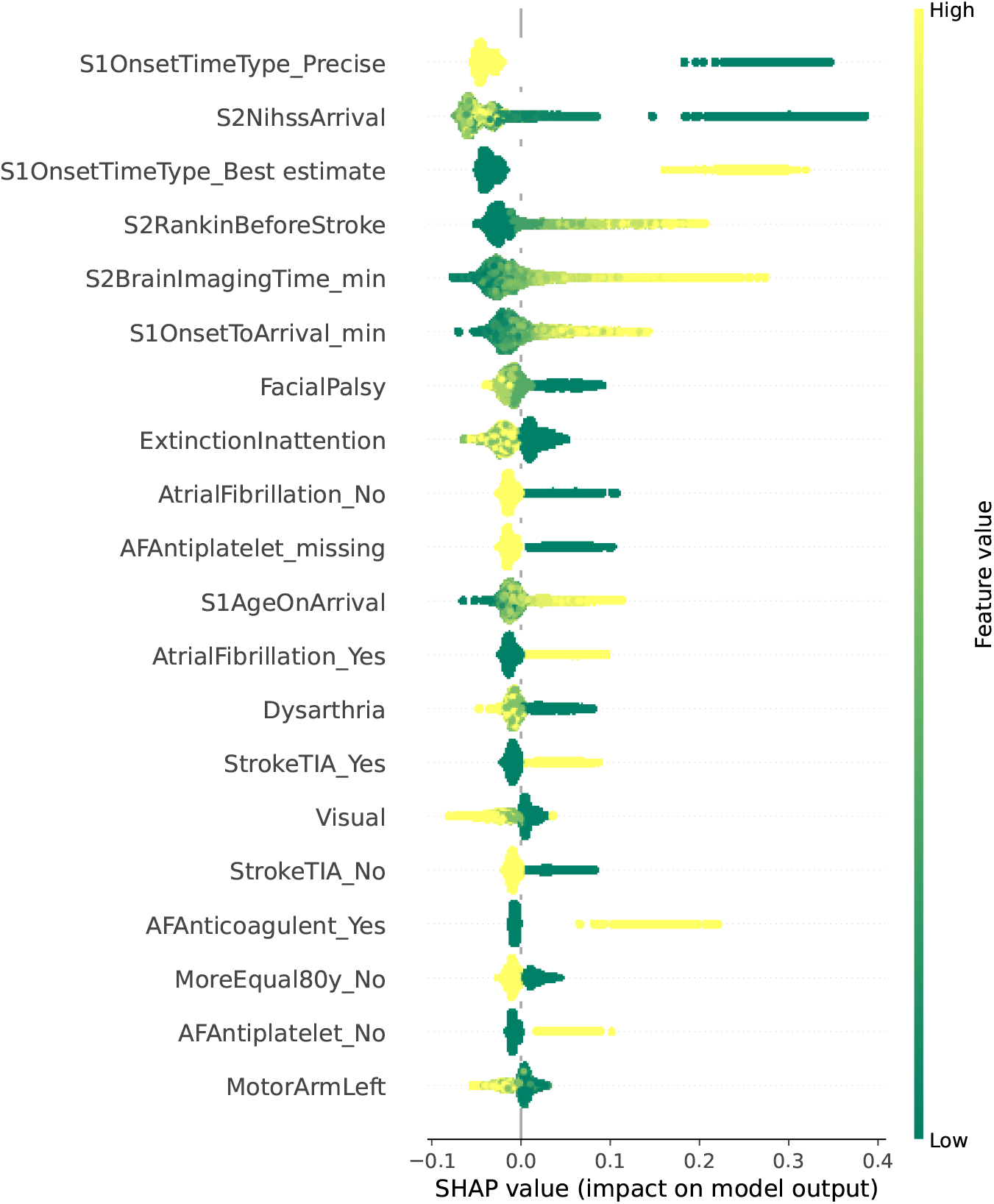
SHAP summary plot for a random forest trained to classify whether a patient would have been thrombolysed at over 70% of hospitals (class 0) or is contentious (class 1).

For example, from Figure 6 the most important feature in determining whether a patient would be thrombolysed, or is contentious, is whether or not the onset time is precise. This is a binary feature, where a low value (ie, 0) has a positive SHAP value, corresponding to the onset time being uncertain and pushing a patient towards the contentious class. In contrast, a high value (ie, 1) represents onset time being precisely known and the corresponding SHAP value is negative, pushing a patient towards the class that over 70% of hospitals would have thrombolysed.

## 4. Discussion

Our methods demonstrate how ML can be used alongside clinical registry data for retrospective audit of decision making. Specifically we have demonstrated that ML can be used to group hospitals according to their decision making and characterise patients that would have been treated differently had they attended a different hospital.

In our example we have used SSNAP, a clinical registry of in-hospital stroke care in the United Kingdom, to investigate variation in thrombolysis use for acute ischemic stroke in patients having a stroke out of hospital. By training separate RFs on patients from each of 132 hospitals, we found the RFs performed well on their own patients (mean accuracy 84.5%, mean AUC 0.90). Using these RFs, we assessed the amount of agreement between hospitals by putting all patients through all models, and we found that hospitals were much more likely to agree on the patients not to thrombolyse than those to thrombolyse. Defining contentious patients as those in which there is less agreement between hospitals in how to treat (30-70% of hospitals would have thrombolysed), we trained an additional RF to discriminate between contentious patients and patients that over 70% of hospitals would have thrombolysed. We found that contentious patients are principally characterized by uncertainty in time of stroke onset (and therefore time since stroke), milder strokes (as measured by the NIHSS) and higher levels of prior disability (as measured by the mRS).

In the United Kingdom the current rate of thrombolysis use is 11.1% of all stroke, which is lower than the target in the NHS Long Term Plan of 20% and there is significant variation between hospitals (0-24.5%). This proportion is also significantly lower than rates reported from large registries in other European countries [21], which suggests that there are significant numbers of stroke patients in the UK who stand to benefit from thrombolysis but who are not being treated. The median proportion of patients in whom the time of stroke onset is precisely known is 31.9%, and in whom it is a ‘best estimate’ is 36.8%, with 31.4% classified with an unknown time of onset (SSNAP Annual Results Portfolio, 2020-21 www.strokeaudit.org). However, there is a remarkable variation between hospitals in these proportions, ranging from 0.4-73.3% in our three-year national sample, a degree of variation that is difficult to understand. The willingness of individual clinicians to estimate the time of onset may relate to their own perception of the balance of risk and benefit from thrombolysis, particularly among patients at either end of the scale of neurological severity and in those with a greater degree of pre-stroke disability. A previous study [15] investigated factors that influence variation in thrombolysis decision-making using a discrete choice experiment involving 138 clinicians, who were presented with hypothetical vignettes and asked how they would treat the patients described. This study also found that stroke severity and pre-stroke disability influenced a clinician’s decision of whether or not to thrombolyse a patient. More recent imaging methods for acute stroke, using CT perfusion or MRI techniques, now offer the prospect of identifying patients eligible for thrombolysis according to a ‘tissue window’ as opposed to the more widely used ‘time window’ [22]. These developments would permit a more objective assessment of the ‘tissue at risk’ as the justification for thrombolysis at any time point after onset (known or unknown), and may enable clinical decision-making to become less variably based on individual clinicians’ judgements about the accuracy of onset times or pre-stroke disabilities. Moving towards greater objectivity in deciding eligibility for thrombolysis may reduce the significant and unexplained variation in practice that we have highlighted in this study, which persists over more than 25 years since the first publication of the evidence for thrombolysis in acute ischemic stroke [23]. Identifying and characterizing the types of patients in whom eligibility is contentious also suggests areas where further clinical research is appropriate, and may help to improve the training of clinical decision-makers in thrombolysis, and thereby increase the consistency in thrombolysis use in the future.

### 4.1. Study Limitations

When investigating the calibration of the hospital models we found that hospitals with both lower thrombolysis rates and fewer patients than average had less well calibrated models. This highlights the fact that when using an ML approach its important to consider whether there is both a sufficient amount of data in total (number of patients) and sufficient data in each class (thrombolysis rate) for the algorithms to perform well.

Performance of ML algorithms is also dependent on the level of detail contained in the data being used. We have demonstrated that hospital models perform well on their own patients, however it is likely that there are parameters that are not measured in SSNAP which are also significant contributors to variation in thrombolysis practice. It is therefore possible that parameters not present in the data, such as organizational factors like the availability and proximity of a CT scanner, or the availability of a stroke physician, impact model performance.

Our predictions about differences in clinical decision-making are necessarily at site level. We pick up on general differences in attitude to thrombolysis between sites, but we cannot detect differences that exist between individual clinicians (as decision-making is the end-result of a process, and may be collective, it may be that decisions can never be fully assigned to an individual).

In the example presented we used random forest algorithms due to their robustness, and previous work [24] has shown that, for this specific example, RF performs comparably to a neural network, and is superior to a logistic regression. However, the methods we have presented are more general and can be used alongside any ML algorithm.

## 5. Conclusion

The use of machine learning as applied to large-scale national stroke registry data can improve our understanding of the sources of variation in decision-making between hospitals in critical areas of clinical practice such as thrombolysis in acute ischemic stroke. The learning derived from advanced analysis could be used to reduce clinical variation through more effective audit and feedback to participating sites, and increase both the consistency of decision-making and the overall use and population benefit from treatments. The wider adoption of machine learning techniques in other comprehensive clinical registries similarly offers the prospect of reducing clinical variation in many other important disease areas.

## Data Availability

The data used in this study was obtained from the Healthcare Quality Improvement Partnership (HQIP) UK

## Acknowledgments

We would like to thank the SAMueL project team (Julia Frost, Kristin Liabo, Kerry Pearn, Tom Monks, Zhivko Zhelev, Stuart Logan, Ken Stein, Leon Farmer, Penny Thompson) for their input into this work.

## References

[1] Richard E Gliklich, Nancy A Dreyer, and Michelle B Leavy. Registries for evaluating patient outcomes: a User’s guide [Internet]. 2014.

[2] Robin Burgess. New principles of best practice in clinical audit. Radcliffe Publishing, 2011.

[3] Sean Ekins, Ana C Puhl, Kimberley M Zorn, Thomas R Lane, Daniel P Russo, Jennifer J Klein, Anthony J Hickey, and Alex M Clark. Exploiting machine learning for end-to-end drug discovery and development. Nature materials, 18(5):435–441, 2019.

[4] Giovanni Cammarota, Gianluca Ianiro, Anna Ahern, Carmine Carbone, Andriy Temko, Marcus J Claesson, Antonio Gasbarrini, and Giampaolo Tortora. Gut microbiome, big data and machine learning to promote precision medicine for cancer. Nature reviews gastroenterology & hepatology, 17(10):635–648, 2020.

[5] Cameron R Olsen, Robert J Mentz, Kevin J Anstrom, David Page, and Priyesh A Patel. Clinical applications of machine learning in the diagnosis, classification, and prediction of heart failure. American Heart Journal, 2020.

[6] Hyunna Lee, Eun-Jae Lee, Sungwon Ham, Han-Bin Lee, Ji Sung Lee, Sun U Kwon, Jong S Kim, Namkug Kim, and Dong-Wha Kang. Machine learning approach to identify stroke within 4.5 hours. Stroke, 51(3):860–866, 2020.

[7] Nathan Peiffer-Smadja, Timothy Miles Rawson, Raheelah Ahmad, Albert Buchard, P Georgiou, F-X Lescure, Gabriel Birgand, and Alison Helen Holmes. Machine learning for clinical decision support in infectious diseases: a narrative review of current applications. Clinical Microbiology and Infection, 26(5):584–595, 2020.

[8] Quinlan D Buchlak, Nazanin Esmaili, Jean-Christophe Leveque, Farrokh Farrokhi, Christine Bennett, Massimo Piccardi, and Rajiv K Sethi. Machine learning applications to clinical decision support in neurosurgery: an artificial intelligence augmented systematic review. Neurosurgical review, 43(5):1235–1253, 2020.

[9] Intercollegiate Stroke Working Party. Sentinel Stroke National Audit Programme (SSNAP). London: Royal College of Physcians, 2015.

[10] Adrian R Parry-Jones, Lizz Paley, Benjamin D Bray, Alex M Hoffman, Martin James, Geoffrey C Cloud, Pippa J Tyrrell, Anthony G Rudd, and SSNAP Collaborative Group. Care-limiting decisions in acute stroke and association with survival: analyses of UK national quality register data. International Journal of Stroke, 11(3):321–331, 2016.

[11] Benjamin D Bray, Lizz Paley, Alex Hoffman, Martin James, Patrick Gompertz, Charles DA Wolfe, Harry Hemingway, Anthony G Rudd, and SSNAP Collaboration. Socioeconomic disparities in first stroke incidence, quality of care, and survival: a nationwide registry-based cohort study of 44 million adults in england. The Lancet Public Health, 3(4):e185–e193, 2018.

[12] Benjamin D Bray, Geoffrey C Cloud, Martin A James, Harry Hemingway, Lizz Paley, Kevin Stewart, Pippa J Tyrrell, Charles DA Wolfe, Anthony G Rudd, and SSNAP collaboration. Weekly variation in health-care quality by day and time of admission: a nationwide, registry-based, prospective cohort study of acute stroke care. The Lancet, 388(10040):170–177, 2016.

[13] Jonathan Emberson, Kennedy R. Lees, Patrick Lyden, Lisa Blackwell, Gregory Albers, Erich Bluhmki, Thomas Brott, Geoff Cohen, Stephen Davis, Geoffrey Donnan, James Grotta, George Howard, Markku Kaste, Masatoshi Koga, Ruediger Von Kummer, Maarten Lansberg, Richard I. Lindley, Gordon Murray, Jean Marc Olivot, Mark Parsons, Barbara Tilley, Danilo Toni, Kazunori Toyoda, Nils Wahlgren, Joanna Wardlaw, William Whiteley, Gregory J. Del Zoppo, Colin Baigent, Peter Sandercock, and Werner Hacke. Effect of treatment delay, age, and stroke severity on the effects of intravenous thrombolysis with alteplase for acute ischaemic stroke: a meta-analysis of individual patient data from randomised trials. The Lancet, 384(9958):1929– 1935, 2014.

[14] Clare R. Carter-Jones. Stroke thrombolysis: Barriers to implementation. International Emergency Nursing, 19(1):53–57, January 2011.

[15] Aoife D. Brún, Darren Flynn, Laura Ternent, Christopher I Price, Helen Rodgers, Gary A Ford, Matthew Rudd, Emily Lancsar, Stephen Simpson, John Teah, and Richard G Thomson. Factors that influence clinicians’ decisions to offer intravenous alteplase in acute ischemic stroke patients with uncertain treatment indication: Results of a discrete choice experiment. International Journal of Stroke, 13(1):74–82, 2018.

[16] thomas Brott, Harold P Adams Jr, Charles P Olinger, John R Marler, William G Barsan, Jose Biller, Judith Spilker, Renee Holleran, Robert Eberle, and Vicki Hertzberg. Measurements of acute cerebral infarction: a clinical examination scale. Stroke, 20(7):864–870, 1989.

[17] Leo Breiman. Random forests. Machine Learning, 45(1):5–32, 2001.

[18] Fabian Pedregosa, Gaël Varoquaux, Alexandre Gramfort, Vincent Michel, Bertrand Thirion, Olivier Grisel, Mathieu Blondel, Peter Prettenhofer, Ron Weiss, Vincent Dubourg, Jake Vanderplas, Alexandre Passos, David Cournapeau, Matthieu Brucher, Matthieu Perrot, and Édouard Duchesnay. Scikit-learn: Machine learning in Python. Journal of Machine Learning Research, 12:2825–2830, 2011.

[19] John C. Platt. Probabilistic Outputs for Support Vector Machines and Comparisons to Regularized Likelihood Methods. In Advance in Large Margin Classifiers, volume 10, pages 61–74. MIT Press, 1999.

[20] Scott M Lundberg and Su-In Lee. A Unified Approach to Interpreting Model Predictions. In I. Guyon, U. von Luxburg, S. Bengio, H. Wallach, R. Fergus, S. Vishwanathan, and R. Garnett, editors, Advances in Neural Information Processing Systems 30, pages 4765–4774. Curran Associates, Inc., 2017.

[21] Laurien S Kuhrij, Michel WJM Wouters, Renske M van den Berg-Vos, Frank-Erik de Leeuw, and Paul J Nederkoorn. The dutch acute stroke audit: benchmarking acute stroke care in the netherlands. European stroke journal, 3(4):361–368, 2018.

[22] Götz Thomalla, Florent Boutitie, Henry Ma, Masatoshi Koga, Peter Ringleb, Lee H Schwamm, Ona Wu, Martin Bendszus, Christopher F Bladin, Bruce CV Campbell, et al. Intravenous alteplase for stroke with unknown time of onset guided by advanced imaging: systematic review and meta-analysis of individual patient data. The Lancet, 396(10262):1574–1584, 2020.

[23] issue Plasminogen Activator for Acute Ischemic Stroke. The national institute of neurological disorders and stroke rt-pa stroke study group. N Engl J Med, 333(24):1581–1587, 1995.

[24] Michael Allen, Charlotte James, Julia Frost, Kristin Liabo, Kerry Pearn, Tom Monks, Zhivko Zhelev, Stuart Logan, Richard Everson, Martin James, and Ken Stein. Stroke Audit Machine Learning (SAMueL): Use of simulation and machine learning to identify key levers for maximising the disability benefit of intravenous thrombolysis in acute stroke pathways. DOI: 10.5281/zenodo.5078131. https://samuel-book.github.io/samuel-1/.

