## Supplementary Material for "Using machine learning and clinical registry data to uncover variation in clinical decision making"

**Supplementary Information**

| Feature | Description | Category |
|---|---|---|
| Stroke Team | Pseudonymised SSNAP team unique identifier | Categorical |
| Pathway | Episode number. Refers to the patient's pathway (i.e. team transfers) | Ordinal |
| Age | Age on arrival aggregated to 5 year bands | Discrete |
| >= 80 years | Whether the patient is >= 80 years old at the moment of the stroke | Binary |
| Gender | Gender | Binary |
| Ethnicity | Patient Ethnicity. Aggregated to White, Black, Mixed, Asian and Other | Categorical |
| Onset in hospital | Whether the patient was already an inpatient at the time of stroke | Binary |
| Onset to arrival (minutes) | Time from symptom onset to arrival at hospital in minutes, where known and if out of hospital stroke | Continuous |
| Onset Date Type | Whether the date of onset given is precise, best estimate or if the stroke occurred while sleep | Categorical |
| Onset Time Type | Whether the time of symptom onset given is precise, best estimate, not known | Categorical |
| Arrive by Ambulance | Whether the patient arrived by ambulance | Binary |
| Admission Hour | Hour of arrival, aggregated to 3 hour epochs | Categorical |
| Admission Day | Day of week at the moment of admission | Categorical |
| Admission Quarter | Year quarter (Q1: Jan-Mar; Q2:April-Jun; Q3: Jul-Sept; Q4: Oct-Dec) | Categorical |
| Admission Year | Year of admission | Categorical |
| Congestive Heart Failure | Comorbidbities: Pre-Stroke Congestive Heart Failure | Binary |
| Hypertension | Comorbidbities: Pre-Stroke Systemic Hypertension | Binary |
| Atrial Fibrillation | Comorbidities: Pre-Stroke Atrial Fibrillation (persistent, permanent, or paroxysmal) | Binary |
| Diabetes | Comorbidities: Pre-Stroke Diabetes Mellitus | Binary |
| Stroke TIA | Comorbidities: Pre-Stroke history of stroke or Transient Ischaemic Attack (TIA) | Binary |
| AF Antiplatelet | Only available if "Yes" to Atrial Fibrillation (Q2.1.3). Whether the patient was on antiplatelet medication prior to admission | Binary |

| Feature | Description | Category |
|---|---|---|
| AF Anticoagulent | Prior to 01-Dec-2017: Only available if "Yes" to Atrial Fibrillation (Q2.1.3); From 01-Dec-2017: available even if patient is not in Atrial Fibrillation prior to admission. Whether the patient was on anticoagulant medication prior to admission | Binary |
| AF Anticoagulent Vitamin K | If the patient was receiving anticoagulant medication, was it vitamin K antagonists | Binary |
| AF Anticoagulent DOAC | If the patient was receiving anticoagulant medication, was it direct oral anticoagulants (DOACs) | Binary |
| AF Anticoagulent Heparin | If the patient was receiving anticoagulant medication, was it Heparin | Binary |
| New AF Diagnosis | Whether a new diagnosis of Atrial Fibrillation was made on admission | Binary |
| Rankin Before Stroke | Patient's modified Rankin Scale score before this stroke (Higher values indicate more disability) | Ordinal |
| Loc | National Institutes of Health Stroke Scale Item 1a Level of Consciousness (higher values indicate more severe deficit) | Ordinal |
| Loc Questions | National Institutes of Health Stroke Scale Item 1b Level of Consciousness Questions (higher values indicate more severe deficit) | Ordinal |
| Loc Commands | National Institutes of Health Stroke Scale Item 1c Level of Consciousness Commands (higher values indicate more severe deficit) | Ordinal |
| Best Gaze | National Institutes of Health Stroke Scale Item 2 Best Gaze (higher values indicate more severe deficit) | Ordinal |
| Visual | National Institutes of Health Stroke Scale Item 3 Visual Fields (higher values indicate more severe deficit) | Ordinal |
| Facial Palsy | National Institutes of Health Stroke Scale Item 4 Facial Paresis (higher values indicate more severe deficit) | Ordinal |
| Motor Arm left | National Institutes of Health Stroke Scale Item 5a Motor Arm - Left (higher values indicate more severe deficit) | Ordinal |
| Motor Arm right | National Institutes of Health Stroke Scale Item 5b Motor Arm - Right (higher values indicate more severe deficit) | Ordinal |

| Feature | Description | Category |
|---|---|---|
| Motor Leg left | National Institutes of Health Stroke Scale Item 6a Motor Leg - Left (higher values indicate more severe deficit) | Ordinal |
| Motor Leg right | National Institutes of Health Stroke Scale Item 6b Motor Leg - Right (higher values indicate more severe deficit) | Ordinal |
| Limb Ataxia | National Institutes of Health Stroke Scale Item 7 Limb Ataxia (higher values indicate more severe deficit) | Ordinal |
| Sensory | National Institutes of Health Stroke Scale Item 8 Sensory (higher values indicate more severe deficit) | Ordinal |
| Best Language | National Institutes of Health Stroke Scale Item 9 Best Language (higher values indicate more severe deficit) | Ordinal |
| Dysarthria | National Institutes of Health Stroke Scale Item 10 Dysarthria (higher values indicate more severe deficit) | Ordinal |
| Extinction Inattention | National Institutes of Health Stroke Scale Item 11 Extinction and Inattention (higher values indicate more severe deficit) | Ordinal |
| NIHSS Arrival | National Institutes of Health Stroke Scale score on arrival at hospital | Discrete |
| Brain Imaging Time (minutes) | Time from Clock Start to brain scan. In minutes. "Clock Start" is used throughout SSNAP reporting to refer to the date and time of arrival at first hospital for newly arrived patients, or to the date and time of symptom onset if patient already in hospital at the time of their stroke. | Continuous |
| Stroke Type | Whether the stroke type was infarction or primary intracerebral haemorrhage | Binary |
| TIA in last month | Whether the patient had a Transient Ischaemic Attack during the last month. Item from the SSNAP comprehensive dataset questions (not mandatory) | Binary |

Table S1: Features in SSNAP used for training the combined model

| Feature | % Missing | Imputation method |
|---|---|---|
| AF Antiplatelet | 79.3 | missing |
| AF Anticoagulent | 50.6 | missing |
| AF Anticoagulent Vitamin K | 62.5 | missing |
| AF Anticoagulent DOAC | 62.5 | missing |
| AF Anticoagulent Heparin | 62.5 | missing |
| New AF Diagnosis | 72.0 | missing |
| Loc Questions | 2.2 | 0 |
| Loc Commands | 2.1 | 0 |
| Best Gaze | 2.7 | 0 |
| Visual | 3.6 | 0 |
| Facial Palsy | 2.1 | 0 |
| Motor Arm left | 2.1 | 0 |
| Motor Arm right | 2.1 | 0 |
| Motor Leg left | 2.2 | 0 |
| Motor Leg right | 2.2 | 0 |
| Limb Ataxia | 4.0 | 0 |
| Sensory | 3.5 | 0 |
| Best Language | 2.3 | 0 |
| Dysarthria | 2.8 | 0 |
| Extinction Inattention | 2.8 | 0 |
| NIHSS Arrival | 5.8 | 0 |
| Brain Imaging Time (minutes) | 0.2 | 9999 |
| Stroke Type | 0.2 | missing |
| TIA in last month | 91.0 | missing |

Table S2: Amounts of missing data in SSNAP and imputation method used.

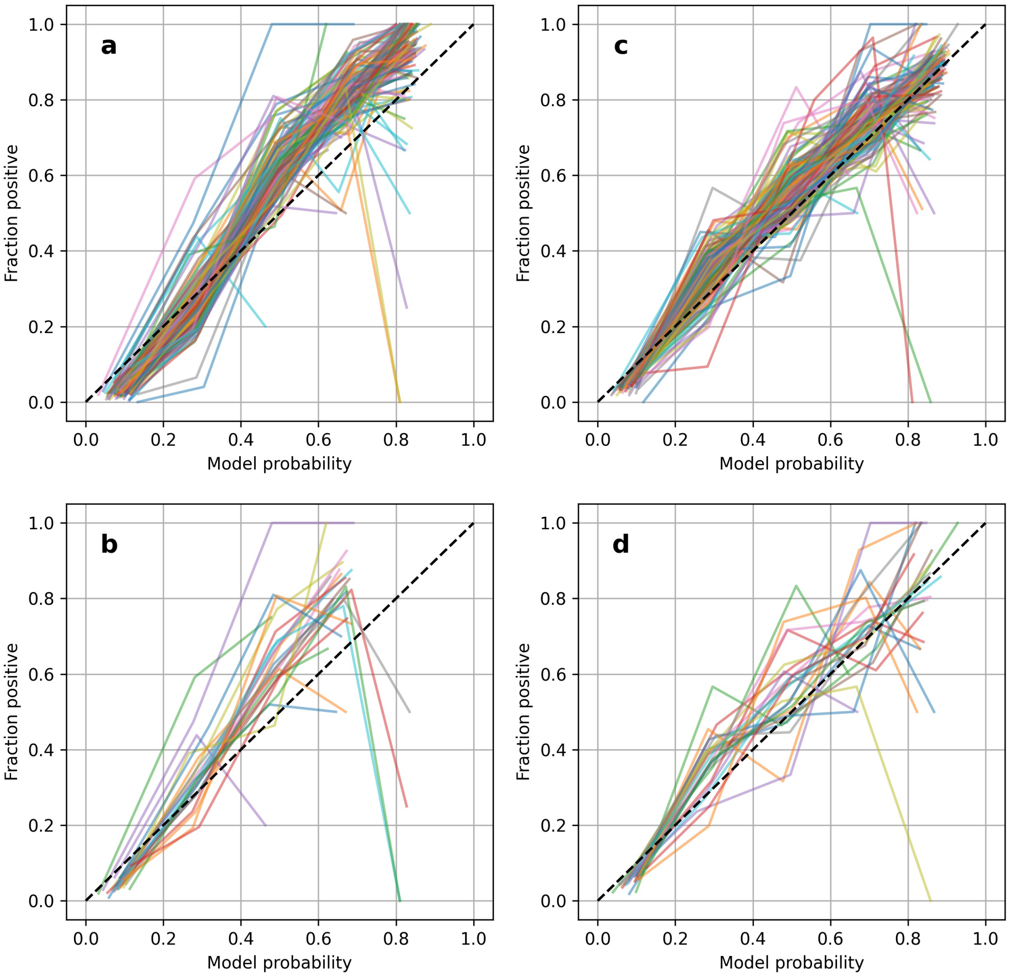


Figure S1: **a** Calibration curves for all hospital models **b** Calibration curves for hospital models that were not well calibrated **c** Calibration curves for all hospital models after applying Platt Scaling **d** Calibration curves for hospitals models that were originally not well calibrated after applying Platt Scaling

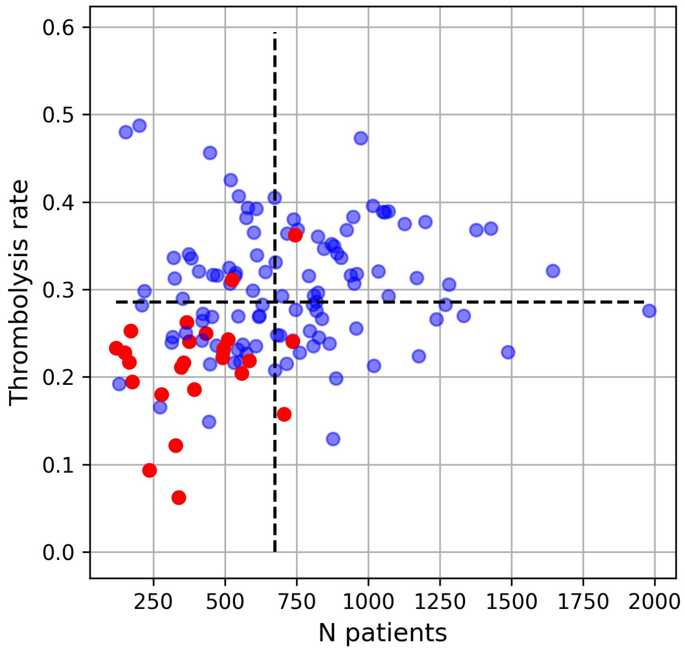


Figure S2: Number of patients (x-axis) vs thrombolysis rate (y-axis) for hospitals that were well calibrated (blue) and those that were not well calibrated (red). Black dashed lines represent mean values along each axis.

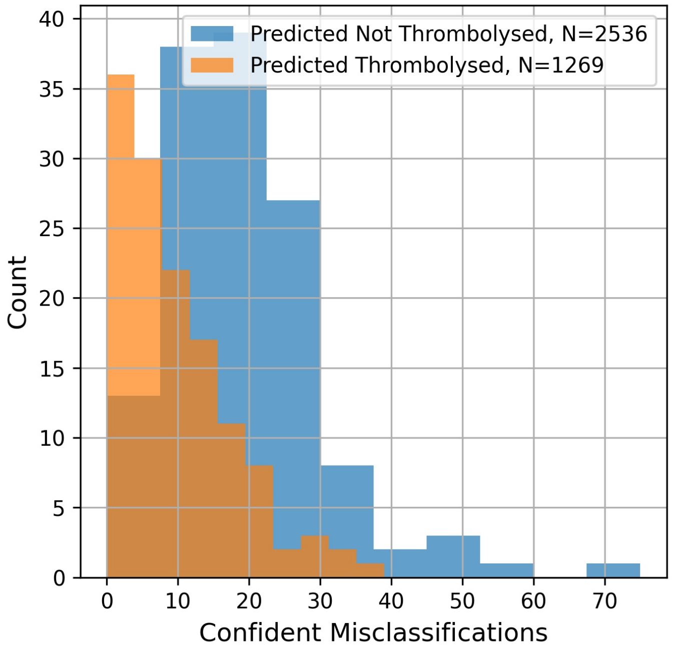


Figure S3: Histogram of confident misclassifications for hospital models, stratified by model outcome. A confident misclassification is defined as a misclassification with model probability $< 0.2$ or $> 0.8$.

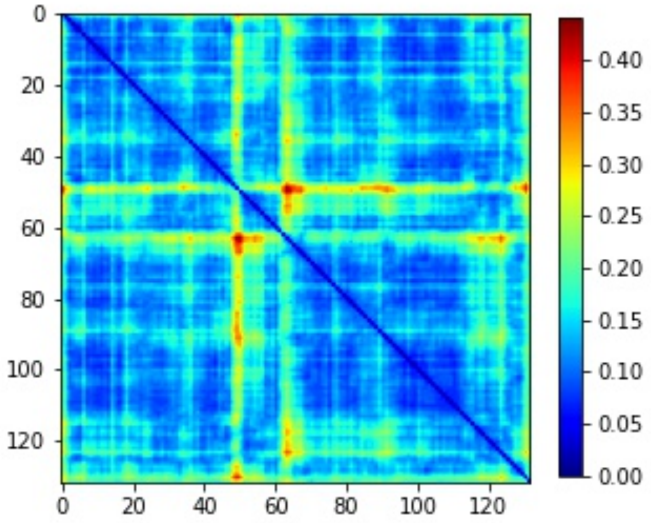


Figure S4: Pairwise Hamming distance between the decisions on treatment of a standard cohort of 10,000 patients made at each hospital.
